# Cohort studies on melanoma and keratinocyte skin cancer: a systematic review

**DOI:** 10.1101/2025.08.13.25332927

**Authors:** Catherine M. Olsen, Rachel E. Neale, David C. Whiteman

## Abstract

The incidence of cutaneous malignancies is increasing worldwide, presenting an important public health burden. Cohort studies can provide high quality data on the epidemiology of these cancers, and are invaluable for deriving measures of disease burden used to inform prevention, diagnosis and treatment. We conducted a systematic review of the literature to summarise the characteristics of cohort studies that have published one or more papers describing the epidemiology of melanoma and/or keratinocyte cancers. Eligible studies were population-based cohort studies that have published findings on incidence or etiology of melanoma or keratinocyte cancer (including associations with phenotypic, environmental, and genetic factors). We excluded clinical cohorts focused on survivorship outcomes. We searched MEDLINE 1950 (U.S. National Library of Medicine, Bethesda, MD, USA), the ISI Science Citation Index (1990 to 31 July 2025) and the reference lists of retrieved articles, imposing no language restrictions. We identified 22 eligible cohort studies, 20 of which had published on melanoma, and 16 on keratinocyte cancer. Nine were conducted in the United States, eleven in Europe, and two in Australia. There was substantial variability in terms of cohort size, risk factor information recorded at baseline, and other data collected (e.g., health services, genetic). Only three studies were specifically designed to examine skin cancers as study endpoints, and only two cohorts pre-specified both melanoma and keratinocyte cancer endpoints. Our summary provides a resource for skin cancer researchers conducting investigations into the causes, burden and prevention of these important cancers.

## INTRODUCTION

The incidence of skin cancer, both cutaneous melanoma and the more common keratinocyte cancers (KC) - basal cell carcinoma (BCC) and squamous cell carcinoma (SCC) - is rising in most light-skinned populations across the world.^1,2^ Strategies to control these cancers include primary prevention and early detection, both of which are dependent on knowledge of the important factors that confer increased risk. To date, most of our knowledge about the risk factors for melanoma and keratinocyte cancers derives from case-control studies, which are prone to selection and recall biases. While population-based prospective cohort studies are not immune to bias, they can mitigate some of these limitations, and offer more robust estimates for causal inference than retrospective study designs. Risk estimates from population-based cohort studies are increasingly used to derive quantitative measures that are used to inform public health policy (e.g. population attributable fractions and potential impact fractions). Furthermore, authoritative international cancer agencies that provide advice regarding strength of association for cancer risk factors place greater emphasis on evidence derived exclusively from prospective studies.^3^

Our aim was to summarise the characteristics of cohort studies that have reported findings on the epidemiology of melanoma and/or keratinocyte cancers.

## METHODS

We conducted a systematic review to identify population-based prospective studies that have published one or more papers on melanoma or keratinocyte cancer. Eligibility criteria for study inclusion were pre-specified, as were the specific data elements to be extracted from each identified cohort study.

### Data sources and searches

Eligible studies published up to July 31, 2025, were identified by searching the MEDLINE 1950 (U.S. National Library of Medicine, Bethesda, MD, USA), the ISI Science Citation Index (1990 to 31 July 2025), and by hand-searching the reference lists of retrieved articles. We did not impose any language restrictions on the search. We did not search for conference abstracts, unpublished studies, or other grey literature.

For computer searches, we used the following medical subject heading (MeSH) terms or text words: “melanoma”, “keratinocyte cancer”, “basal cell carcinoma”, “BCC”, “squamous cell carcinoma”, “SCC”, “cohort study”, “prospective”, “longitudinal”. The search terms are provided in Supplementary Methods.

We read the abstracts of all identified studies to exclude those that were clearly not relevant, and read the full texts of the remaining articles to determine if they met the study inclusion criteria.

### Inclusion and exclusion criteria

We included population-based cohort studies that have published on incidence or aetiology of melanoma or keratinocyte cancer (including associations with phenotypic, environmental, and genetic factors).

### Data extraction

A single reviewer (CO) abstracted data from identified studies using a standardized data abstraction form. The following information was recorded: cohort name; geographic location; numbers of participants; sex (male/female); years of follow-up; number of cases (melanoma/KC; taken from the most recent report); baseline assessment of phenotype (pigmentation factors etc.), nevi, sun exposure and sun protection; case definition (i.e., histological confirmation); availability of pathology data; linkage to health services and costings data; collection of biological specimens; and whether repeatability or validation studies had been conducted.

### Data synthesis and analysis

We conducted a qualitative synthesis of the identified cohort studies.

## RESULTS

We found 1176 published reports from which we identified 22 relevant cohort studies (Figure 1): 20 had published at least one paper on melanoma^4–22^ (Table 1) and 16 at least one paper on keratinocyte cancer^4,7,10,12,14,16,19–21,23–28^ (Table 2) (14 cohorts had published at least one paper on both outcomes^4,6,7,10,12,14,16,19–21,25–28^).

**Table 1.** Characteristics of population-based cohort studies that have published data on melanoma.

| COHORT | ACRONYM | COHORT SIZE | FOLLOW-UP | SEX | GEOGRAPHIC LOCATION | NUMBER OF CASES | PHENOTYPE AT BASELINE | NEVI | UV/SUN EXPOSURE | SUN PROTECTION | HISTOLOGICAL CONFIRMATION OF CASES | PATHOLOGY | BIOLOGICAL SPECIMENS | HEALTH SERVICE DATA | COSTING DATA | REPEATABILITY/ VALIDITY |
| --- | --- | --- | --- | --- | --- | --- | --- | --- | --- | --- | --- | --- | --- | --- | --- | --- |
| NURSES HEALTH STUDY (I AND II)<sup>4,30</sup> | NHS | NHS I: 86,380<br>NHS II: 104,100 | 1976-present | Female | US | 2439 up to 2011/2012 for NHS/HPFS | 1982 survey for NHS, 1991 survey for NHS II | 1982 survey for NHS, 1991 survey for NHS II | Average July ambient erythematous UV radiation | Not at baseline | Self-reported, confirmed by medical records | No | 200,000 participants across all three cohorts | No | No | Not reported |
| HEALTH PROFESSIONALS FOLLOW-UP STUDY<sup>4</sup> | HPFS | 46,934 | 1986-present | Male | US | 2439 up to 2011/2012 for NHS/HPFS | 1988 survey | 1988 survey | Average July ambient erythematous UV radiation | Not at baseline | Self-reported, confirmed by medical records | No | 200,000 participants across all three cohorts | No | No | Not reported |
| MELANOMA INQUIRY OF SOUTHERN SWEDEN<sup>5</sup> | MISS | 40,000 | 1990-present | Female | Sweden | 155 invasive, 60 *in situ* | Yes | Yes | Yes | Yes | Yes | No | DNA collection began in 2011 | No | No | Yes (Westerdahl et al., 1996<sup>31</sup>) |
| NORWEGIAN WOMEN AND CANCER STUDY<sup>6,25</sup> | NOWAC | 165,772 | 1991-present | Female | Norway | 1774 | Yes | Yes | Yes | Yes | Yes | No | Blood collected for 60,000/170,000 women | No | No | Yes (Veierod et al., 2008<sup>32</sup>) |
| FRENCH TEACHERS COHORT<sup>7,33</sup> | E3N | 100,000 | 1992-present | Female | France | 611 | Yes | Yes | In a nested-case control study only (2008) | In a nested-case control study only (2008) | Yes | No | 47,000 saliva samples; not all genotyped | No | No | Yes |
| EUROPEAN PROSPECTIVE INVESTIGATION INTO CANCER AND NUTRITION<sup>8,34</sup> | EPIC | 519,978 | 1992-present | Both | Europe | 4372 | No | No | No | No | Yes | No | Blood collected from 385,747/519,978 | No | No | Yes (diet only) |
| UNITED STATES RADIOLOGIC TECHNOLOGISTS COHORT<sup>9,35</sup> | USRT | 68,588 | 1983-present | Both (78.8% female) | US | 837 | No (second survey) | No | Yes | No | Self-reported; some validated through medical record review | No | No | No | No | Not reported |
| JANUS COHORT<sup>10</sup> |  | 318,628 | 1972-present | Both | Norway | 2570 | No | No | Yes - 1972-1991 only, ambient | No | Yes | No | Yes - serum | No | No | Not reported |
| THE MULTIETHNIC COHORT<sup>11</sup> |  | 215,251 | 1993-present | Both | US | 581 invasive, 412 *in situ* | Yes | No | No | No | Yes | No | Blood/urine on 70,000/215,000 collected in 2001-2005 | No | No | Yes (diet only) |
| THE NAMBOUR SKIN CANCER PREVENTION TRIAL<sup>12</sup> |  | 1,621 | 1992-2006 | Both | Australia | 33 | Yes | Yes | Yes | Yes | Yes | No | No | No | No | Yes (diet only) |
| CANCER PREVENTION STUDY-3 COHORT<sup>13</sup> | CPS-3 | 39,368 | 2006-present | Both | US | 60 | Not reported | Not reported | Not reported | Not reported | Self-reported in 2011; registry linkage confirmed 42/60 |  | Blood | No | No | Not reported |
| THE DIET, CANCER AND HEALTH COHORT<sup>14</sup> |  | 160,725 | 1993-present | Both | Denmark | 357 | Yes | Yes | No | No | Yes | No | Not reported | No | No | Not reported |
| NIH-AARP DIET AND HEALTH STUDY<sup>15,36</sup> | NIH-AARP | 447,357 | 1995-present | Both | US | 5034 invasive, 3284 *in situ* | No | No | Yes | No | Yes | No | Not reported | No | No | Not reported |
| THE MILLION WOMENS' COHORT<sup>16</sup> | MWS | 1.32 million | 1996-present | Female | UK | Not reported | No (12-year survey subset) | No | No | No | Yes | No | 5% sample only (blood) | Yes | ? | Yes |
| VITAMINS AND LIFESTYLE (VITAL) COHORT STUDY<sup>17</sup> | VITAL | 77,738 | 2000-present | Both (48% male) | US | 309 invasive, 257 *in situ* | Yes | Yes | No | No | Yes | No | No | No | No | Yes (diet only) |
| CANCER PREVENTION STUDY II (CPS-II) AND CPS-II NUTRITION COHORT<sup>18</sup> | CSP-II | 1.2 million | 1982-present | Both | US | 1238 | No | No | No | No | Yes | No | No | No | No | Not reported |
| NORWEGIAN-SWEDISH WOMEN'S LIFESTYLE AND HEALTH COHORT STUDY/WOMEN'S LIFESTYLE AND HEALTH COHORT STUDY<sup>19</sup> |  | 106,379 | 1991-2005 | Female | Norway, Sweden | 412 | Yes | Yes | Yes | No | Yes | No | No | No | No | Not reported |
| WOMEN'S HEALTH INITIATIVE<sup>20,37</sup> | WHI | 59,575 | 1993-present | Female | US | 1154 | Yes | No | Not at baseline - year 4 survey | Not at baseline - year 4 survey | Yes | No | No | No | No | Yes (diet and other characteristics; not phenotype or sun exposure) |
| QSKIN SUN AND HEALTH STUDY<sup>21,38</sup> | QSKIN | 43,794 | 2010-present | Both | Australia | 706 invasive and 1423 *in situ* | Yes | Yes | Yes | Yes | Yes | Yes | Saliva collected on 17,965 participants; all genotyped | Yes | Yes | Yes (Morze et al., 2012<sup>39</sup>; Mortimore et al, 2021<sup>40</sup>) |
| UK BIOBANK<sup>22</sup> | UKB | 500,000 | 2006-2010 to present | Both | UK | 1592 | Yes | Yes | Yes | No | No | No | Blood, saliva, urine; all genotyped | Yes | No | Yes (some phenotypes – see Schoeler et al., 2025<sup>41</sup>) |

**Table 2.** Characteristics of population-based cohort studies that have published data on keratinocyte cancers.

| COHORT | ACRONYM | COHORT SIZE | FOLLOW-UP | SEX | GEOGRAPHIC LOCATION | NUMBER OF CASES | PHENOTYPE AT BASELINE | NEVI | UV/SUN EXPOSURE | SUN PROTECTION | HISTOLOGICAL CONFIRMATION OF CASES | PATHOLOGY | BIOLOGICAL SPECIMENS | HEALTH SERVICE DATA | COSTING DATA | REPEATABILITY/ VALIDITY |
| --- | --- | --- | --- | --- | --- | --- | --- | --- | --- | --- | --- | --- | --- | --- | --- | --- |
| WOMEN'S HEALTH INITIATIVE<sup>20,37</sup> | WHI | 59,575 | 1993-present | Female | US | 9593 KC (not differentiated) | Yes | No | Not at baseline - year 4 survey | Not at baseline - year 4 survey | No | No | No | No | No | Yes (diet and other characteristics; not phenotype or sun exposure) |
| THE NAMBOUR SKIN CANCER PREVENTION TRIAL<sup>12</sup> |  | 1,621 | 1992-2006 | Both | Australia | 281 BCC; 188 SCC | Yes | Yes | Yes | Yes | Yes | No | No | No | No | Yes (diet only) |
| NURSES HEALTH STUDY (I AND II)<sup>4,30</sup> | NHS | NHS I: 86,380<br>NHS II: 104,100 | 1976-present | Female | US | 17,556 BCC, 2233 SCC | 1982 survey for NHS, 1991 survey for NHS II | 1982 survey for NHS, 1991 survey for NHS II | Average July ambient erythematous UV radiation | Not at baseline | Self-reported, confirmed by medical records | No | 200,000 participants across all three cohorts | No | No | Not reported |
| HEALTH PROFESSIONALS FOLLOW-UP STUDY<sup>4</sup> | HPFS | 46,934 | 1986-present | Male | US | 13,092 BCC, 1756 SCC | 1988 survey | 1988 survey | Average July ambient erythematous UV radiation | Not at baseline | Self-reported, confirmed by medical records | No | 200,000 participants across all three cohorts | No | No | Not reported |
| THE DIET, CANCER AND HEALTH COHORT<sup>14</sup> |  | 160,725 | 1993-present | Both | Denmark | 3,465 BCC, 341 SCC | Yes | Yes | No | No | Yes | No | Not reported | No | No | Not reported |
| THE MILLION WOMENS' COHORT<sup>16</sup> | MWS | 1.32 million | 1996-present | Female | UK | 48,666 BCC; 6699 SCC | No (12 year survey subset) | No | No | No | Yes | No | 5% sample only (blood) | Yes | No | Yes |
| FRENCH TEACHERS COHORT<sup>7,33</sup> | E3N | 100,000 | 1992-present | Female | France | 2426 BCC; 451 SCC | Yes | Yes | In a nested-case control study only (2008) | In a nested-case control study only (2008) | Yes | No | 47,000 saliva samples; not all genotyped | No | No | Yes |
| SUPPLÉMENTATION EN VITAMINES ET MINÉRAUX ANTIOXYDANTS (SU.VI.MAX) COHORT<sup>23</sup> | SU.VI.MAX | 5880 | 1994-2007 | Both | France | 106 BCC, 18 SCC | Yes | No | Yes | Yes | Yes | No | Blood sample for 1760/13.017 trial participants | No | No | Yes (diet only) |
| NORWEGIAN-SWEDISH WOMEN'S LIFESTYLE AND HEALTH COHORT STUDY<sup>19</sup> |  | 106,379 | 1991-2009 | Female | Norway, Sweden | 141 SCC | Yes | Yes | Yes | No | Yes | No | No | No | No | Not reported |
| JANUS COHORT<sup>10</sup> |  | 318,628 | 1972-present | Both | Norway | 1191 BCC, 633 SCC | No | No | Yes - 1972-1991 only, ambient | No | Yes | No | Yes - serum | No | No | Not reported |
| ROTTERDAM STUDY<sup>24</sup> |  | 15,000 | 1990-present | Both | Holland | 1528 BCC | Yes | No | Yes | Yes | Yes (GWAS) |  | No | Yes | No | No |
| QSKIN SUN AND HEALTH STUDY<sup>21,42</sup> | QSKIN | 43,794 | 2010-present | Both | Australia | 640 BCC 193 SCC 316 unknown | Yes | Yes | Yes | Yes | Yes | Yes | Saliva collected on 17,965 participants; all genotyped | Yes | Yes | Yes (Morze et al., 2012<sup>39</sup>; Mortimore et al, 2021<sup>40</sup>) |
| NORWEGIAN WOMEN AND CANCER STUDY<sup>25</sup> | NOWAC | 172,472 | 1991-present | Female | Norway | 871 | Yes | Yes | Yes | Yes | Yes | No | Blood collected for 60,000/170,000 women | No | No | Yes (Veierod et al., 2008<sup>32</sup>) |
| UNITED STATES RADIOLOGIC TECHNOLOGISTS COHORT<sup>26</sup> | USRT | 62,595 | 1972-present | Both | US | 6,339 BCC 1253 SCC | No (second survey) | No | Yes | No | Self-reported; some validated through medical record review | No | No | No | No | Not reported |
| THE MULTIETHNIC COHORT<sup>27</sup> |  | 215,251 | 1993-present | Both | US | 265 SCC | Yes | No | No | No | Yes | No | Blood/urine on 70,000/215,000 collected in 2001-2005 | No | No | Yes (diet only) |
| UK BIOBANK<sup>28</sup> | UKB | 500,000 | 2006-2010 to present | Both | UK | 11,762 BCC<br>1623 SCC | Yes | Yes | Yes | No | No | No | Blood, saliva, urine; all genotyped | Yes | No | Yes (some phenotypes – see Schoeler et al., 2025<sup>41</sup>) |

**Figure 1.**
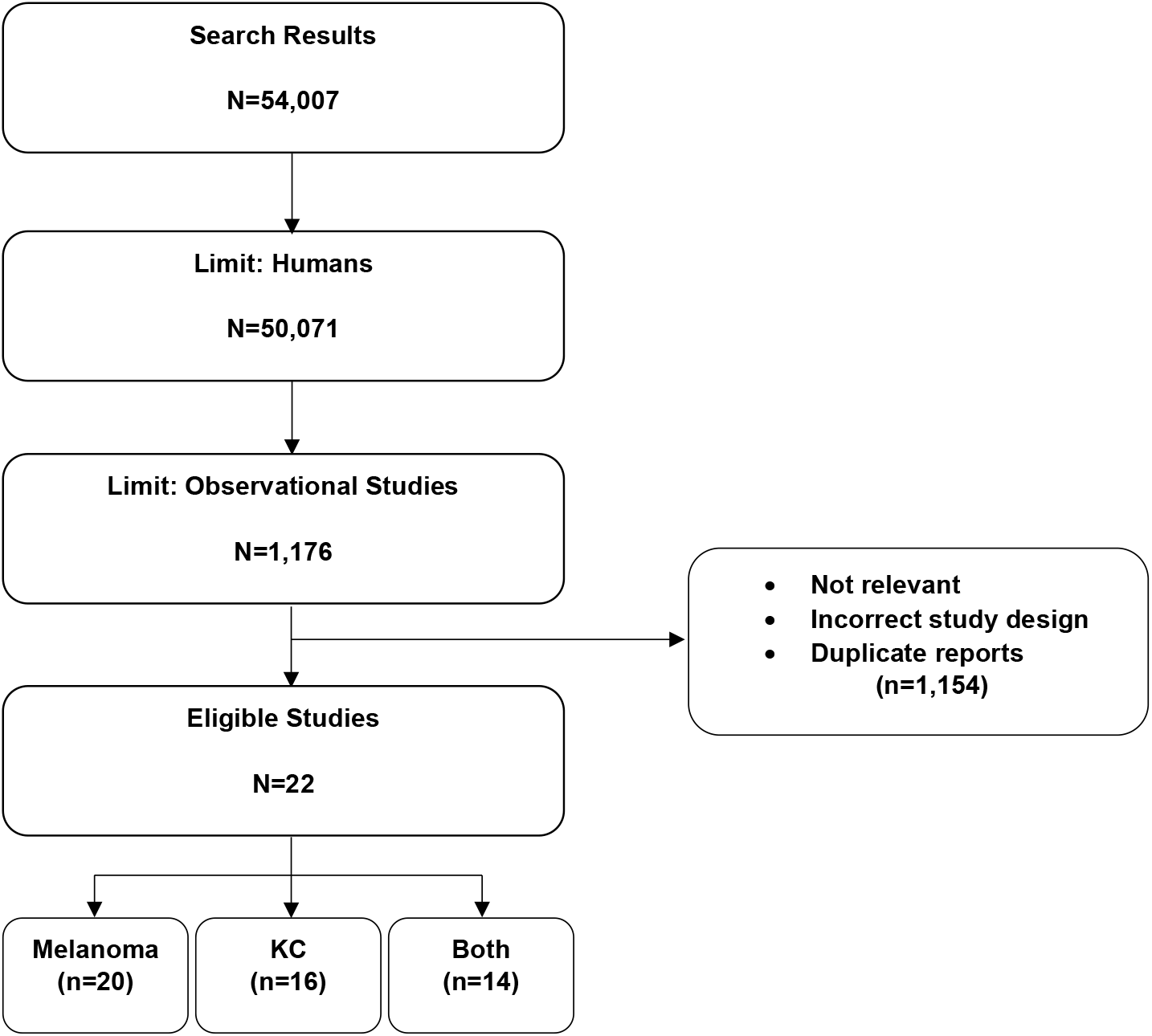
Flowchart for study inclusion.

Of the 20 cohorts that published on melanoma, 12 included both men and women; seven included women only and the remaining cohort included men only. Nine were conducted in the United States, nine in Europe, and two in Australia (Table 1). Of the 16 cohorts that have published on KC, nine included both men and women; six included women only and the remaining cohort included men only. Five were conducted in the US, nine in Europe and two in Australia (Table 2). Cohort size ranged from less than 2000 to 1.32 million, and the number of cases of melanoma ranged from 33 to 5034 (BCC from to 106 to 17,556, and SCC from 18 to 2233).

Histological confirmation of cases (melanoma/KC) was ascertained in most of the cohorts. Biological samples suitable for genetic analyses were collected in approximately half of the cohort studies, mostly for a sub-set of the full cohort.

Only three of the cohorts were established specifically to investigate skin cancer endpoints;^5,12,21^ the remaining cohorts did not capture important aspects of phenotype (e.g., skin type, nevi, hair color, tanning ability) and/or environmental risk (e.g., sun exposure, sun protection) at baseline. The three purpose-designed skin cancer cohorts were the Nambour Study (n=1621),^12^ MISS (Melanoma Inquiry in Southern Sweden, n=29,508, women only)^5^ and the QSkin Sun and Health Study (QSkin; n=43,794).^21^ Both the Nambour Study and MISS collected comprehensive baseline data on risk factors, but because of either small size (Nambour) or low incidence (Swedish women), they have very few melanoma events despite long follow-up (Nambour = 33 melanomas; MISS = 155 invasive, 60 *in situ*). Only the Nambour study and the QSkin cohort have data on both melanoma and KC as study endpoints. Neither Nambour nor MISS have linked their cohorts to other datasets to capture health services events or health costings data, and neither has published melanoma genotype data. The QSkin cohort reported 706 invasive and 1423 *in* situ melanoma cases after a median of 11.4 years, and is the only cohort study with linked health services data as well as genotype data.

## DISCUSSION

Cohort studies are powerful tools for epidemiologic discovery, and for developing public health measures and practices;^29^ their impact on public health is unquestioned. They have distinct advantages over case-control studies by characterizing exposures and risk factors prior to disease onset, which reduces important biases. They are also valuable for understanding the genetic basis of complex diseases. The breadth and reliability of prospectively ascertained environmental exposure data also allows the examination of potentially important and clinically relevant gene-environment interactions.

In this systematic review of the literature, we identified 22 population-based cohorts that published on the incidence or etiology of melanoma and/or KC, of which 20 reported on melanoma and 16 on KC. BCCs and SCCs are not registered in many parts of the world, so prospective data for these cancers is scarcer than for melanoma.

Most of the cohort studies identified were not established specifically to examine skin cancer outcomes and thus did not collect information about important risk factors and potential confounding factors at baseline. Conversely, the three studies that were purposely designed to investigate skin cancer collected tailored information at baseline to meet the study goals. ^5,12,21^ Two of these cohorts can provide value through the examination of all skin cancer outcomes simulataneously.^12,21^ Notwithstanding their primary focus on cancers of the skin, all three of those ‘specialized’ cohorts can also provide valuable insights for other disease endpoints, particularly those that might share risk factors.

This study is novel in systematically summarizing the characteristics of all population-based cohort studies with data on melanoma and KCs. We believe we have identified all informative reports from prospective cohort studies that have published on skin cancer outcomes. We specifically excluded clinical cohorts of skin cancer patients established to assess survivorship and other clinical outcomes; similarly, we excluded other selective cohorts that imposed restrictive exclusion criteria on participants that limited generalizability. Our review highlights the small number of cohorts that have collected comprehensive skin cancer risk factor information in the correct temporal sequence, and as such provides a resource for skin cancer researchers conducting investigations into the causes, burden and prevention of these important cancers.

## Data Availability

Not applicable - this is a systematic review (without meta-analysis) and all included studies have been published.

