## Supplementary Material for "Cohort studies on melanoma and keratinocyte skin cancer: a systematic review"

### PubMed Search

(cohort study OR prospective OR longitudinal) AND (skin cancer OR melanoma OR keratinocyte cancer OR BCC or basal cell carcinoma OR squamous cell carcinoma OR SCC) NOT (head and neck OR oesophageal squamous OR esophageal squamous OR lung cancer OR larynx)

(("cohort studies"[MeSH Terms] OR ("cohort"[All Fields] AND "studies"[All Fields]) OR "cohort studies"[All Fields] OR ("cohort"[All Fields] AND "study"[All Fields]) OR "cohort study"[All Fields]) OR ("longitudinal studies"[MeSH Terms] OR ("longitudinal"[All Fields] AND "studies"[All Fields]) OR "longitudinal studies"[All Fields] OR "prospective"[All Fields]) OR longitudinal[All Fields]) AND (("skin neoplasms"[MeSH Terms] OR ("skin"[All Fields] AND "neoplasms"[All Fields]) OR "skin neoplasms"[All Fields] OR ("skin"[All Fields] AND "cancer"[All Fields]) OR "skin cancer"[All Fields]) OR ("melanoma"[MeSH Terms] OR "melanoma"[All Fields]) OR (("keratinocytes"[MeSH Terms] OR "keratinocytes"[All Fields] OR "keratinocyte"[All Fields]) AND ("neoplasms"[MeSH Terms] OR "neoplasms"[All Fields] OR "cancer"[All Fields])) OR BCC[All Fields] OR ("carcinoma, basal cell"[MeSH Terms] OR ("carcinoma"[All Fields] AND "basal"[All Fields] AND "cell"[All Fields]) OR "basal cell carcinoma"[All Fields] OR ("basal"[All Fields] AND "cell"[All Fields] AND "carcinoma"[All Fields])) OR ("carcinoma, squamous cell"[MeSH Terms] OR ("carcinoma"[All Fields] AND "squamous"[All Fields] AND "cell"[All Fields]) OR "squamous cell carcinoma"[All Fields] OR ("squamous"[All Fields] AND "cell"[All Fields] AND "carcinoma"[All Fields])) OR SCC[All Fields]) NOT (("Head Neck"[Journal] OR ("head"[All Fields] AND "and"[All Fields] AND "neck"[All Fields]) OR "head and neck"[All Fields]) OR (oesophageal[All Fields] AND squamous[All Fields]) OR (esophageal[All Fields] AND squamous[All Fields]) OR ("lung neoplasms"[MeSH Terms] OR ("lung"[All Fields] AND "neoplasms"[All Fields]) OR "lung neoplasms"[All Fields] OR ("lung"[All Fields] AND "cancer"[All Fields]) OR "lung cancer"[All Fields]) OR ("larynx"[MeSH Terms] OR "larynx"[All Fields]))

N=54,007

Limit Humans:

N=50,071

Limit: Observational Studies

N=1176
